# Prediction of COVID-19 Diagnosis from Healthy and Pneumonia CT scans using Convolutional Neural Networks

**DOI:** 10.1101/2022.10.20.22281334

**Authors:** Rushil Srirambhatla, Helmet T. Karim

## Abstract

**Background:** Current methods of COVID-19 detection from other respiratory illnesses using computed tomography (CT) scans are highly inaccurate. However, understanding pathogen-specific immune responses can help reduce inconsistencies and improve the accuracy of COVID-19 and Pneumonia detection. A deep learning model using Relief-based feature selection (RBAs) was developed to detect COVID-19 and Pneumonia. Patient-specific Class Activation Maps (CAMs) were produced to highlight immunopathogenic differences and identify differences between COVID-19 and Pneumonia on CT scans.

**Methods:** To examine the effect on lung lesions, a COVIDx CT-2 dataset, containing CT scans from 3,745 patients, was examined. We developed an algorithm to convert the 3-D CT scan of each patient into multiple 2-D slices. Altogether, there were 194,344 2-D slices retrieved from 3,745 CT Scans. The distribution of slices was 67%-20%-17% consisting of COVID-19, Pneumonia, and normal CT scan, respectively. An AlexNet architecture was implemented with additional feature extraction layers (containing RBA) and classification layers to perform deep learning. The 2-D slices were divided into 3 groups: Training, Test, and Validation. The training set consisted of 70% of the data, the test set consisted of 20% of the data, and the validation consisted of 10% of the data. After training, unique CAMs were generated on patient CT scans using the immunopathogenic differences to highlight COVID-19 and Pneumonia related abnormalities.

**Results:** The model accurately distinguished hyperinflammation in COVID-19 patients from Pneumonia patients and achieved a validation accuracy of 95.60% and a false-positive rate of 4.65%. Additionally, the segmented lung, shown by the patient-specific CAMs, identified higher levels of inflammation in the lung of COVID scans compared to the other two groups.

**Discussion:** The use of deep learning in disease diagnosis and prevention has provided many avenues to advance current techniques. Likewise, in this analysis, deep learning was shown to successfully predict COVID-19 via CT scan. By providing patient-specific CAMs, the model can be used to not just aid in diagnosis but potentially also to evaluate serial chest CT scans for treatment.

## Introduction

Severe acute respiratory syndrome coronavirus 2 (SARS-CoV-2) caused by COVID-19 (Coronavirus disease 2019), is an acute respiratory illness that has caused over 5.69 million deaths worldwide (Bener 2022 et al). This illness is characterized by inflamed and irritated respiratory tracts, and in severe cases, affects the alveoli present in both lungs. While reverse transcription-polymerase chain reaction (RT PCR) has been the most commonly used method for detecting COVID-19, computed tomography (CT) scans have been used, albeit variably, by medical professionals to aid in diagnosis in some scenarios especially early on. During the initial stages of the pandemic, CT scans were used to determine if continued isolation was required. Additionally, it has been reported that CT aided in revealing abnormal findings in over 80% of hospitalized patients and contributed to implementing effective treatment plans for them (Mølhave 2021 et al).

Some tests have high false negative rates and current processes of detecting COVID-19 from a CT scan require huge amounts of time and resources (Pecoraro 2021 et al). CT scans have a much more specialized role in care and are often not used except for cases in hospitals in some scenarios. However, they still play a role in treatment guidance for severe cases. The average screening time for CT scans ranges between 5-30 minutes (Cleveland Clinic, 2020), this places a significant burden on medical professionals given the rising number of cases and reduced healthcare workforce. Furthermore, differences between COVID-19 and pneumonia can make diagnosis less clear on CT scans. Developing approaches that improve the speed and accuracy of these approaches as well as more clearly help assess severity can potentially improve diagnosis, management, and treatment primarily in hospitalized patients by identifying critical features of the CT.

Deep learning has the potential to quickly read CT scans and identify areas of focus for healthcare workers. Using these approaches, we can develop class activation maps or identify areas in a single image that the algorithms are utilizing to predict diagnosis. We identified a publicly available dataset of CT scans and trained a deep-learning model to detect COVID-19 and pneumonia from chest CT scans and provide patient-specific class activation maps (CAMs). This allows the identification of regions of lungs with high vulnerability and track the management of those specific abnormalities or be used to assess the severity of the diagnosis. Additionally, CAMs with critical features in chest CT were generated to help show inflammation status that may otherwise be overlooked through human observation.

## Methods

### Data

Deep learning was performed by utilizing data from the COVIDx CT-2A (Gunraj, 2022). COVIDx CT-2A contains 194,344 2-D slices from 3,745 patients’ CT scans. This dataset has been collected from 7 different initiatives: (1) China National Center for Bioinformation (CNCB) (China), (2) National Institutes of Health Intramural Targeted Anti-COVID-19 (ITAC) Program (countries unlisted), (3) Negin Radiology Medical Center (Iran), (4) Union Hospital and Liyuan Hospital of Huazhong University of Science and Technology (China), (5) COVID-19 CT Lung and Infection Segmentation initiative, annotated and verified by Nanjing Drum Tower Hospital (Iran, Italy, Turkey, Ukraine, Belgium, some countries unknown), (6) Lung Image Database Consortium (LIDC) and Image Database Resource Initiative (IDRI) (USA), and (7) Radiopaedia collection (Iran, Italy, Australia, Afghanistan, Scotland, Lebanon, England, Algeria, Peru, Azerbaijan, some countries unknown). The mean patient age of 51 years with a standard deviation of 16.7 years. The data consisted of a 67%, 20%, and 17% split of COVID-19, pneumonia, and normal CT scan, respectively. Figure 1 outlines the workflow for the model.

**Figure 1.**
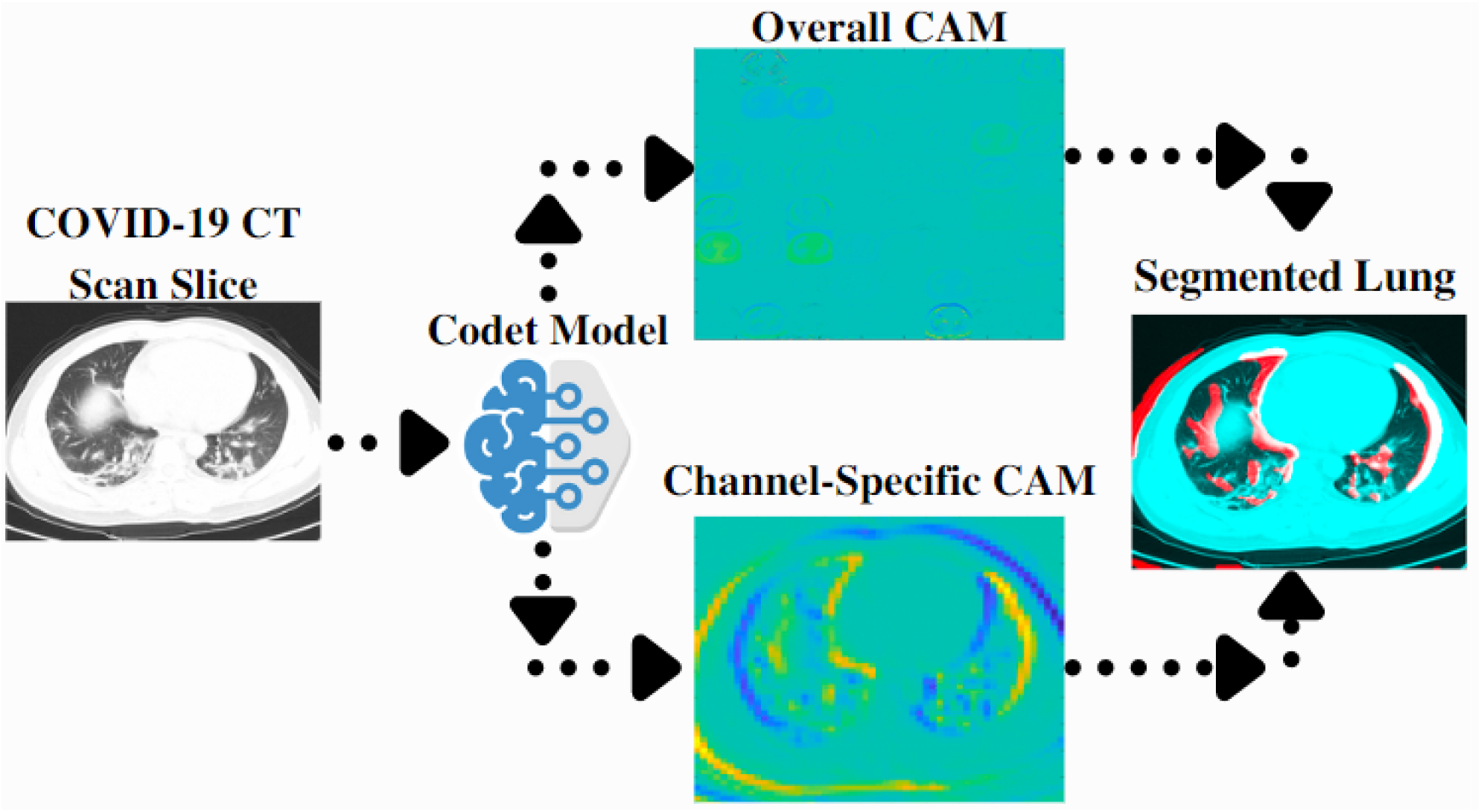
Model workflow - data was split into train, test, and validation sets. The model was trained and patient-specific CAMs were generated. We developed a covid detection model that was able to generate a CAM that could be segmented to identify regions of importance.

### Data Preprocessing

The COVIDx CT-2A dataset was used and was imported into MATLAB – where data pre-processing, deep learning model architecture construction, and model training and validation was conducted. The 194,344 2-D slices were augmented in 2 ways. First, imlocalbrighten function was used to increase the brightness of the slices, making pathogen-specific features more prominent. Second, imrotate was used to orient all slices in the same direction (180 degrees), for better feature extraction and deep learning. The data was divided into a training set (70%), a test set (20%), and a validation set (10%), to effectively measure the model’s accuracy.

### Architecture Construction and Hyperparameter Tuning

The model was initially validated under two pre-trained networks: ResNet-50 and AlexNet. AlexNet performed better than ResNet-50 during training, however, so its architecture was further altered to focus on feature extraction. The AlexNet architecture was modified in two ways. Two Fully Connected Layers and ReLU layers were added for better data compilation from previous layers. Initial Learn Rate was reduced to 0.0001, and Mini Batch Size was decreased to 16 for increased feature extraction through Relief Based Algorithms.

### Model Training and Validation

The model with Relief Based Algorithms was used to train, validate, and predict diagnosis. We generated a confusion matrix to calculate the accuracy, sensitivity, specificity, false positive rate, false negative rate, positive predictive value (PPV), and negative predictive value (NPV) in the validation set to gauge performance.

### Class Activation Maps

Patient-specific CAMs were generated after model training, using the RBA feature selection, to segment the lung. This was through a 3-step process. First, with the use of RBAs, one activation map from the Convolutional Layer in the model architecture was selected. Second, imgaussfilt was applied to this image, with a Kernel value of 5, to filter and smooth the image. Next, the pixel intensities (where 0.55 > pixel intensities > 0.35) collected were utilized to threshold the filtered image to isolate areas of high predictive value. The threshold image was closed using the imclose function and a structural element of a disk with a radius of 1. This closed image was overlaid using imfuse on the original 2-D slice to observe areas of importance.

## Results

During training, the mini-batch loss decreased from 0.37 to 0.01, and the training accuracy reached 100%. On the validation set, the model exhibited a 95.6% accuracy. On the validation set, the model reached a sensitivity of 95.6% and a specificity of 95.6%. An algorithm that identifies key distinct features using linear binary patterns (LBPs) was used to quantify the differences between the 3 classes. Specifically, the algorithm resulted in a low LBP value range between 2 Covid-19 CT scans of 0.0027, and a high LBP value range between a Covid and a Non-Covid CT scan of 0.1373. Furthermore, the model exhibited a false positive rate of 4.7%, a positive predictive value of 95.3%, and a negative predictive value of 95.8%.

Relief-based algorithms were used to extract critical features from that CT scan that would distinguish a COVID-19 CT scan from the other two groups. This feature selection method identifies the nearest hit class (closest CT scan to a COVID scan) and the nearest miss class (closest CT scan to a Pneumonia scan). The differences in these features are then computed using algorithms to provide an accurate classification of slices. Using the model-trained features, patient-specific CAMs are generated that segment the lung from chest CT scans and identify areas of importance in red. The model-generated CAMs showcased high levels of inflammation in COVID patients and low levels of inflammation in pneumonia patients (see Figure 2).

**Figure 2.**
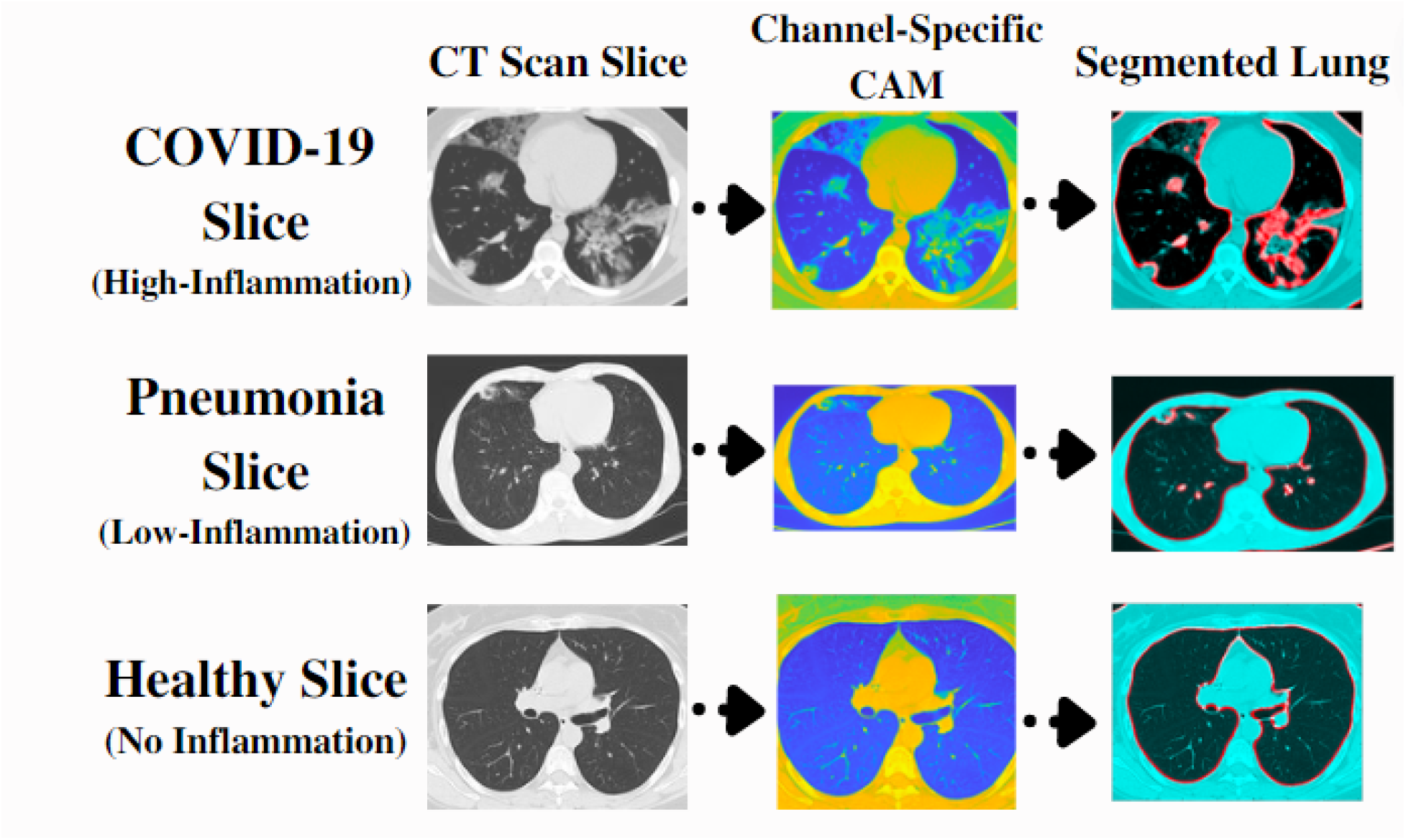
CAMs for one slice of a patient with COVID vs. Pneumonia vs. Healthy. Red areas indicate greater importance compared in rightmost images.

## Discussion

Our model was able to achieve relatively high accuracy, sensitivity, and specificity. We were able to generate patient-specific CAMs that highlight important features of interest in the CT scans. This measures up to past algorithms, delivering high accuracy and sensitivity. Considering the current stress and pressure on healthcare workers, deep learning models may help alleviate such problems by identifying key features of interest and assessing initial diagnostics that could aid physicians.

There have been several deep-learning approaches taken to solve the problem of misdiagnosis. In one study they used a comparative learning approach to reduce overparameterization of the data, as well as implement an incremental augmentation algorithm that exhibits an accuracy of 85.1% to 94.4% accuracy on publicly available datasets. Another study used transfer learning on VGG16, InceptionResNetV2, VGG19, Xception, DenceNet121, MobileNet, and InceptionV3 networks using a large dataset of X-ray images. Their model achieved an accuracy of 99% and a sensitivity of 98.3%. Though the model was able to successfully distinguish radiology images, this was done on a more limited dataset. Another study used the same dataset as used in this model and aimed to improve on previous research studies by expanding the dataset to include patients from different regions and from varying ages. They proposed a lightweight deep neural network that achieves an accuracy of 99.0%, and they extend their scope of the study by understanding the model’s decision-making through GSInquire. We replicate their findings using our model and highlight similar patient-specific CAMs.

We were able to generate detailed patient-specific class activation maps (CAMs) that highlight areas of high inflammation. These CAMs may aid radiologists to detect inflammatory features in CT scans, especially during periods of high burden, healthcare worker shortages, and boundary cases with less inflammation. This potentially could be used to develop patient-specific treatment plans based on the inflammatory levels shown by the specific CAMs. For instance, seeing a large overview of inflammation visually may help physicians better assess overall severity.

Patients are often misdiagnosed especially if a CT scan is required. Due to incorrect diagnoses, patients undergo ineffective or unnecessary treatment plans that affect their physical and mental health. This approach may help alleviate some of this burden and aid in diagnostic challenges.

Future studies may investigate clinical utility by evaluating multiple CT scans in the course of treatment. These models provide a demonstration of utility for prediction, but the use of these approaches clinically needs further research to identify critical gaps and barriers.

## Data Availability

All data is available online at https://www.kaggle.com/datasets/hgunraj/covidxct

## Acknowledgments

We would like to thank the organizations and initiatives that collected and shared this data.

